# A Modeling Framework for Evaluating the Synergistic Impact of Structural Interventions on Related Diseases: HIV and Cervical Cancer as Case Study

**DOI:** 10.1101/2025.07.24.25332100

**Authors:** Xinmeng Zhao, Chaitra Gopalappa

## Abstract

**Background:** Women with HIV face elevated cervical cancer risks, compounded by social conditions that influence both disease outcomes. Current models fail to adequately capture the complex interactions between diseases and social determinants.

**Methods:** We enhanced a mixed agent and compartment model for HIV and cervical cancer (MAC-HIV-CC) to model disparities by social conditions. We analyzed the impact of hypothetical 100% efficacious interventions over 30 years (2018-2048): (1) an HIV care intervention that eliminates disparities in viral load suppression between social groups, (2) a sexual behavior intervention aligning behaviors of women who exchange sex with those who do not, and (3) a combination of both interventions.

**Results:** The HIV care intervention reduced HIV incidence by 26.9% and cervical cancer cases by 14.5% among HIV-positive women. The sexual behavior intervention decreased HIV prevalence by 8.1% and HPV prevalence by 36.1% among HIV-positive women engaged in exchange sex. The combination intervention reduced HIV prevalence by 25.3%, HIV incidence by 34.3%, and cervical cancer cases by 37.5% in the target population.

**Conclusions:** The proposed framework provides a novel approach for health equity analyses by modeling social determinants that are common pathways to interrelated diseases and health disparities. Such a model is of significance for cost-effectiveness intervention analyses of interrelated diseases.

## 1. Introduction

In the United States, cervical cancer remains a pressing health concern, with around 13,820 women expected to be diagnosed and approximately 4,360 deaths anticipated annually in 2024 ^1^. Despite the overall decline in incidence and mortality rates ^2^—attributed to advancements in screening, treatment, and the introduction of the HPV vaccine—these benefits have not been uniformly realized across all demographics ^3–5^. This situation is more pronounced among women living with HIV, about 270,000 women in the United States ^6^, whose risk of cervical cancer is 2 to 7 times greater than those without HIV ^7–9^. The heightened risk of cervical cancer in women living with HIV can be partially attributed to the biological risk from a compromised immune system because of HIV, and partially due to the common sexual behavioral mechanisms for HIV and HPV infection ^10^. These biological and behavioral factors significantly increases their vulnerability to acquiring and maintaining HPV infections (2 to 6 times ^11–13^), which in turn, accelerates the progression to cervical cancers ^14–16^.

Further compounding this issue, social conditions significantly influence risky behaviors associated with HIV, HPV, and cervical cancer, thus complicating the management of these diseases. For example, material hardship and unstable housing are closely correlated with higher instances of condomless sex and an increased number of sexual partners ^17,18^, which contribute to the transmission of both HIV and HPV. Additionally, socioeconomic factors such as residing in high Social Vulnerability Index (SVI) areas ^19^, having an annual household income of ≤ $50,000 ^20^, or lacking health insurance ^20^ are linked with lower adherence to cervical cancer screening guidelines, leading to delayed diagnosis of cervical cancer. These same socioeconomic factors also affect HIV management, where individuals facing poverty ^21^ or unstable housing ^21,22^, and lacking insurance ^21,23^, frequently fail to initiate antiretroviral therapy (ART) or achieve viral load suppression (VLS), thereby worsening their health outcomes and potentially increasing HIV transmission. In 2021, it was reported that among those diagnosed with HIV in the US, approximately 41% experience disabilities, 39% are unemployed, 38% live below the poverty threshold, and 17% are homeless ^24^. Similarly, in the literature on cervical cancer, the absence of insurance ^25^ and living in rural areas ^26^ are associated with a greater burden of the disease, underlining the profound impact of social conditions on health outcomes.

Acknowledging social determinants as pathways to health risk behaviors and thus health disparities, Healthy People 2030 ^27^ emphasizes the importance of implementing structural interventions (SI), such as expanding health care coverage, subsidized housing and food programs, improving access to mental healthcare, to mitigate the impact of social conditions on disease transmission and management ^28–31^. Structural interventions are thus integral to increase the impact of behavioral interventions aimed at preventing sexually transmitted diseases (STDs).

While the significance of structural interventions (SI) is clear, challenges persist in effectively informing resource allocation decisions. Traditional models for cost-effectiveness analyses take a disease-centered, typically a single disease, approach to intervention analyses. This method fails to capture the complex interactions between diseases and the broader impact of SIs on overall disease prevention, leading to potentially inaccurate estimations of their cost-effectiveness. For example, current cervical cancer and HIV models do not incorporate social conditions when assessing the impact of HPV vaccination ^32–34^, cervical cancer screening programs ^32,34–36^, and HIV related interventions ^34,37^ on disease outcomes.

Moreover, models that do address social conditions often do not simulate multiple diseases simultaneously or lack detailed mechanistic insights. Typically, statistical models draw data from observational studies and explore the relationship between structural interventions and changes in risky behaviors or direct health outcomes. However, these often provide static snapshots and lack dynamic behavior representation ^38–42^. Current mechanistic models frequently rely on simplified assumptions ^43,44^ or focus on a single disease ^45,46^. This gap highlights the need for models that can integrate multiple infectious diseases and common social drivers.

The objective of this study is to develop a simulation modeling framework for conducting joint structural intervention analyses, particularly in the context of HIV, HPV, and cervical cancer due to their biological and behavioral interactions. This framework is specifically designed to model behaviors by social condition status, enhancing the model’s existing capacity of simulating infections and disease burden as functions of behaviors. Current literature are limited in data on the effectiveness of structural interventions in reducing disparities across social status. Therefore, we evaluated the impact of two hypothetical interventions that were 100% effective is eliminating the disparities across social conditions in health risk behaviors - one focused on HIV care behaviors and another on sexual behaviors among women who exchange sex. While hypothetical, these analyses provide a valuable proof-of-concept. We believe this framework will help steer future research by supporting cost-effectiveness analysis of structural interventions through joint analyses of interconnected diseases and informing public health strategies that address both biological and social determinants of diseases. Additionally, it can direct future data collection on social conditions and behaviors and the impact of structural intervention, leading to more accurate analysis.

## 2. Methodology

In this work, we extended a recently developed novel mixed agent and compartment model for HIV and cervical cancer (MAC-HIV-CC). The MAC-HIV-CC model is a calibrated and validated simulation tool that captures the national-level epidemiology of HIV and HPV infection, as well as the progression to cervical cancer in the U.S ^10^. It employs a newly developed simulation framework suitable for jointly modeling diseases with varying prevalence^47^.

Simulating diseases like HIV, HPV and cervical cancer in a large population such as the U.S. presents significant computational challenges ^10^. While the compartmental model is well-suited for higher-prevalence diseases like HPV, it cannot capture the heterogeneity and network dynamics essential for modeling slower-spreading diseases like HIV. On the other hand, while agent-based network modeling can represent these complexities they struggle to scale efficiently to the national level for lower-prevalence diseases such as HIV and cervical cancer ^6,7^.

The MAC framework overcomes these challenges by adopting a hybrid approach. It simulates persons with at least one lower prevalence or slower spreading disease, in this case, HIV, and their immediate contacts in an agent-based network model. It simulates all other persons, including individuals with only higher-prevalence diseases, such as HPV and its progression to cervical cancer, in a compartmental model. This allows for an efficient simulation of both disease types while maintaining the necessary detail for network dynamics. The framework further utilizes an evolving contact network algorithm (ECNA) ^48^ to maintain partnership dynamics and network dynamics across the compartmental model and the network model.

The following sections will briefly discuss the model’s key components, its calibration and validation, and then detail how the model was extended to incorporate social conditions and the hypothetical scenario analysis we conducted.

### 2.1 Overview of MAC-HIV-CC

The MAC-HIV-CC model monitors HIV-infected persons and their immediate contacts within a dynamic network, tracking changes over time as new infections occur and contacts are added. Each person in the network is characterized by specific attributes, including age, transmission risk group, number of lifetime partners (degree), health status along the HIV related disease and care continuum, and the disease stage of HPV and cervical cancer. Immediate contacts are defined as all the sexual partners an individual could have over their lifetime, who can be either susceptible to or infected with HIV.

The rest of the population - those who are HIV-negative or not immediate contacts of HIV-infected persons - is modeled in a compartment model with differential equations. This compartmental model categorizes individuals by age group, transmission group, degree bin (where degrees are grouped similar to age-groups) and HPV and cervical cancer disease and care stages. Although events related to HIV transmission and progression are modeled at the individual level in the network, the compartmental model also tracks them at the aggregated levels, including CD4 count categories (≤200, 201-350, 351-500, >500) to simplify modeling HPV transmissions and progressions in the overall population. The CD4 count categories, a measure of immune suppression from HIV, helps account for the varying risk of HPV transmission and progression to cervical cancer.

As HIV-susceptible contacts become infected in the network model, their immediate contacts are added into the network. An evolving network algorithm maintains the network dynamics by determining the attributes of partners to newly add, such as age, degree and transmission risk group. Partner attributes tend to be correlated ^49^, and this correlation guides the selection process in the model, ensuring that the evolving contact network accurately reflects the real-world dynamics of disease transmission.

#### Modules in the MAC-HIV-CC Model

The overall epidemiological, demographical, and network dynamics are maintained through simulation of four main modules in the MAC-HIV-CC include:

- **Compartmental Module**: manages demographic features (births, aging, and natural mortality) and simulates HPV disease transmission and cervical cancer progression using differential equations. This module determines HPV disease transmission and cervical cancer progression for both HIV-susceptible and HIV-infected individuals, accounting for the differences in risks based on levels of immune suppression, i.e, CD4 count.
- **Network Transmission Module**: Determines HIV transmission for each individual using a Bernoulli transmission equation based on individual-level sexual behavior, such as number of active sexual partnerships, number of sexual acts and other transmission risk factors.
- **ECNA Module**: Manages the dynamics of partnerships—formation and dissolution—in both the compartmental and network models using sub-algorithms developed from machine learning, stochastic processes, and optimization. These algorithms assign attributes to new contacts of newly HIV-infected individuals to match observed data on age mixing, transmission risk group mixing, and degree mixing. The HPV and cervical cancer disease stages of these newly added individuals are then determined based on the distributions associated with their age, degree, and transmission risk group.
- **Network Disease Module**: Updates the individual-level status of HIV-infected persons, including HIV-specific parameters: CD4 count, viral load, opportunistic infection (OI) incidence, and onset of acquired immune deficiency syndrome (AIDs), changes in HIV diagnosis, care and treatment status, as well as updates in HPV transmission and progression to cervical cancer determined through the compartmental module.

#### Model Calibration and Validation

The MAC-HIV-CC model integrates both HIV and cervical cancer components, each calibrated and validated using extensive national data. The HIV model, based on the validated PATH 4.0 framework ^50^, simulates HIV transmission among heterosexual females (HETF), heterosexual males (HETM), and men who have sex with men (MSM). It utilizes data from national surveillance systems like the National HIV Surveillance System (NHSS), Medical Monitoring Project (MMP), and other significant surveys to ensure accurate representation and validation against epidemic and network features from 2006 to 2017 ^51–56^, encompassing demographical, sexual behavioral, clinical, and HIV care and treatment aspects.

The cervical cancer model simulates high risk HPV and cervical cancer progression in a national population, with sexual behavioral data harmonized with the HIV model. Calibration targets include HPV genotype frequencies and prevalence data by normal cytology and precancer stages from the literature ^57^. Validation involves comparing natural history and status-quo cervical cancer incidence and mortality with registry data ^58,59^, ensuring a robust fit to observed metrics. The model’s details and validation procedures have been presented elsewhere ^10^.

Modeling was performed using NetLogo version 6.4.0 for the agent-based model and Python version 3.11.5 for the compartment model and interfaced with Netlogo using PyNetLogo.

### 2.2. Integration of Social Conditions

Both the compartmental and agent-based components of the MAC-HIV-HPV model were adjusted to model behaviors (*𝓑*) by social status (*𝓢*), as discussed below, prior to modeling infections as functions of behaviors.

#### Social Conditions (𝓢)

Let set {*s*_1_, *s*_2_,…, *s_n_*} represent a set of social conditions (or intermediary, e.g., exchange sex) that may influence individual behaviors and health outcomes. In our model, we considered the following social conditions:

- **Poverty (***s*_1_): Modeled as a binary variable where *s*_1_ = 0 indicates individuals above the federal poverty level, and *s*_1_ = 1 indicates individuals below the federal poverty level.
- **Insurance Status (***s*_2_): Represented as a binary variable with *s*_2_ = 0 for insured individuals and *s*_2_ = 1 for uninsured individuals.
- **Housing Status (***s*_3_): *s*_3_ = 0 denotes stable housing, while *s*_3_ = 1 represents homelessness or unstable housing.
- **Neighborhood (***s*_4_): Defined based on socioeconomic factors, specifically the povery level of the census tract, with *s*_4_ = 0 indicating individuals residing in census tracts where less than 20% of the population is below the federal poverty level, representing favorable neighborhood conditions; *s*_4_ = 1 indicating individuals residing in census tracts where 20% or more of the population is below the federal poverty level, representing disadvantaged neighborhood conditions.
- **Employment (***s*_5_): *s*_5_ = 0 for employed individuals and s_5_ = 1 for unemployed individuals.
- **Education (***s*_6_): *s*_6_ = 0 for individuals with at least a high school degree, and *s*_6_ = 1 for those without a high school degree.
- **Depression (***s*_7_): Depression often co-occurs with other adverse social conditions, so we treated it as part of social conditions here, where *s*_7_ = 0 for individuals with depression, and *s*_6_ = 1 for those without depression.
- **Engagement in Exchange Sex (***s*_8_): Engaging in exchange sex is an intermediary mechanism between social conditions and behavior, i.e., persons who exchange sex have higher number of partners, and social conditions influence participation in exchange sex Here, *s*_8_ = 0 indicates individuals not engaged in exchange sex, and *s*_8_ = 1 indicates those who are.

#### Behaviors (𝓑)

Let set 𝓑 = {*b*_1_, *b*_2_,…, *b*_m_} represent a set of behaviors including sexual practices, disease specific care, or other health-related actions. The behaviors considered in our model include:

- **Number of Sexual Partners (***b*_1_): Represented as categorical variables using ’degree bins’. Each bin corresponds to ranges based on powers of two (e.g., 2^k^ for k = 0, 1, … , 7). This binning approach categorizes individuals into ranges such as 1–2, 3–4, 5–8, up to 128 partners. We do not use a uniform range because degree distribution follows a power-law distribution.
- **Condom Use (***b*_2_): Modeled as a binary variable where *b*_2_ = 0 indicates condom use, and *b*_2_ = 1 indicates no condom use.
- **Other behaviors** (*b_i_*): Depending on the scenario, additional behaviors such as HIV care behavior, vaccination uptake, and cancer screening participation can be incorporated.

In MAC-HIV-HPV model, we added an additional feature *S*, representing a unique social condition combination, along with the original features such as age (*A*), risk group (*R*), degree bin (*D*), and health (*H*) states. That is, instead of modeling each of the *n* social condition as a bivariate variable, we convert it into one categorical variable taking values between 0 and 2*^n^* - 1, where *S* = 0 represents a socially good status, and *S* > 0 implies at least one socially disadvantaged condition. Thus, the compartmental model is a multidimensional array of size *A× R × D × H × S*, and in the network, each agent has features *A, R, D, H, S*.

If *Pr*(*S*), i.e., the joint probability distribution of social conditions, and *Pr*(*B* = *b_i_*I *S*) , i.e.., behaviors conditional to social status are available, we can model behaviors by social status. However, data for such a multivariate joint distribution are not available. Data are typically available as bivariate or pairwise associations between a behavior and a social condition. To address this limitation, our recent work developed a method combining copula and probabilistic graphical modeling to estimate the multivariate joint distribution using the bivariate associations^45^.

We utilized the outcomes of this previous work, specifically the joint distributions, for jointly parametrizing social conditions and behaviors in both the compartmental model and the network as follows. As noted above, the compartmental model was additionally stratified by *S*, and in the network each agent had an additional feature S. In both compartmental model and the network, during the disease transmission phase, behaviors such as condom use were determined by conditional probability of *P*(*b*_2_|*S*). In the disease progression phase, behaviors like care engagement were determined by conditional probability of *P*(*b*_i_|*S*).

### 2.3 Scenario analysis

In the absence of data on effectiveness of structural interventions, to demonstrate the type of analyses that can be conducted through this model, we evaluated the impact of hypothetical interventions that are 100% effective. The first analysis focused on HIV care behaviors among diagnosed HIV individuals, while the second examined sexual behaviors among women who exchange sex.

#### HIV Care Behavior Intervention

We selected viral load suppression (VLS) (*b*_3_) as a proxy for HIV care behavior as it collectively encapsulates linkage to care upon HIV diagnoses, retention in care, and consistent use of ART. That is, achieving VLS indicates that a person is regularly attending medical appointments, adhering to their ART regimen, and effectively managing their HIV. This not only reduces the individual’s viral load to undetectable levels—significantly lowering the risk of HIV transmission—but also stabilizes the immune system, thereby reducing the risk of opportunistic infections, including HPV, and the progression to cervical cancer ^60^.

We focused on seven social conditions (*s*_i_,n = 7), known to be strongly associated with VLS in the literature: poverty (*s*_1_), insurance (*s*_2_), housing status (*s*_3_), neighborhood (*s*_4_), employment (*s*_5_), education (*s*_6_) and depression (*s*_7_). Each social condition was represented by a binary variable, where *s_i_* = 1 indicates disadvantaged social conditions and *s*_i_ = 0 indicates a ^45^ using a copula-informed favorable status. Estimates of *P*(*b*_3_|*s*) were calculated in prior work using a copula-informed MRF model. While this prior work focused on a single disease HIV ^45^, here, we extended it to our MAC-HIV-HPV model. Joint distribution estimates indicate that *P*(*VLS* = 1|*S* > 0) > Pr(VL*S* = 1|*S* = 0), i.e., a larger probability of no viral suppression among persons with atleast one social vulnerability compared to persons with no social vulnerability.

To evaluate the impact of closing these disparity gaps in care, we modeled a hypothetical 100% efficacious structural intervention by modifying the care behaviors of individuals with *S* > 0 to become equal to behaviors in persons with = 0 , i.e., *P*(*b*_3_ = 1|*S* > 0) → *Pr*(*b*_3_ = 1|*S* = 0). We then ran the simulation to assess the impact of this intervention on HPV, HIV, and cervical cancer outcomes.

#### Sexual Behavior Intervention

Our analysis of sexual behavior concentrates on number of sexual partnerships (*b*_1_) and condom usage (*b*_2_). We selected engagement in exchange sex as the primary high-risk social condition, as data on other high-risk populations were limited. Nonetheless, this population encompass a significant proportion of individuals involved in high-risk sexual behaviors compared to other social conditions (Table S3) and is thus among the most vulnerable populations. In our model, engagement in exchange sex is represented as a binary social condition variable, denoted as *s*_8_, where *s*_8_ = 0 indicates individuals not engaged in exchange sex, and *s*_8_ = 1 indicates those engaged in exchange sex.

We examined how engagement in exchange sex influences high-risk behaviors by modeling the conditional probabilities *P*(*s*_8_|*b*_1_) and *P*(*b*_2_|*s*_8_). Since the number of sexual partnerships (*b*_1_) is a categorical variable represented using ‘degree bins’, we utilized the conditional probability distribution *P*(*s*_8_|*b*_1_ = k) for each degree bin k.

To evaluate the impact of closing the disparity gaps influencing sexual behaviors, we modeled a hypothetical 100% efficacious intervention by modifying the behavior in persons who exchange sex to align with those not engaged in exchange sex, i.e., *P*(*s*_8_ = 1|*b*_1_ = k) → *P*(*s*_8_ = 0|*b*_1_ = k) and *P*(*b*_2_ = 1|*s*_8_ = 1) → *Pr*(*b*_2_ = 1|*s*_8_ = 0). We then ran the simulation to assess the impact of this intervention on HPV, HIV, and cervical cancer outcomes.

#### Combination of HIV Care Behavior and Sexual Behavior Intervention

To evaluate the impact of closing the disparity gaps influencing both HIV care behavior and sexual behavior on disease outcomes, we modeled a scenario that included both interventions described above.

#### Simulation Setup and Metrics Collected

We initiated our simulation in 2006 and simulated HIV, HPV, and cervical cancer transmissions and progression over the period 2006 to 2017, calibrating to surveillance data on the coverage of care, treatment, and vaccination for this period. As noted earlier, the calibration and validation of this model was conducted in previous work ^10^. Here, we simulated the model for a period of 30 year, 2018 to 2048, under each of the 3 scenarios described above, and a baseline scenario that assumed care and sexual behavior would be held at the 2018 values.

From each simulation run, we extracted the following metrics: HIV incidence, HIV prevalence, HPV prevalence, and cumulative new cervical cancer cases by HIV status. In this study, we define new cervical cancer cases as transitions from the final precancer stage to the first (undiagnosed) cancer stage, rather than newly diagnosed cases. This approach allows us to isolate the impact of interventions on disease progression without the confounding effects of cancer screening and treatment interventions. Each scenario was simulated 10 times, using the same random seed to calculate pairwise percentage changes for each metric. The percentage change was calculated as

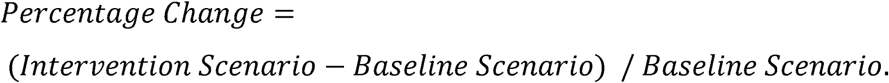

## 3. Results

### HIV Care Behavior

The proportion of VLS among women living with HIV in the ‘baseline’ scenario was approximately 54.6%. A 100% efficacious structural intervention would increase it to 71.5%. This will generate a cumulative (2018-2048) HIV incidence reduction of 26.9% (range 23.5% to 28.2%) (Figure 1a) compared to the baseline scenario, in the heterosexual female group.

**Figure 1.**
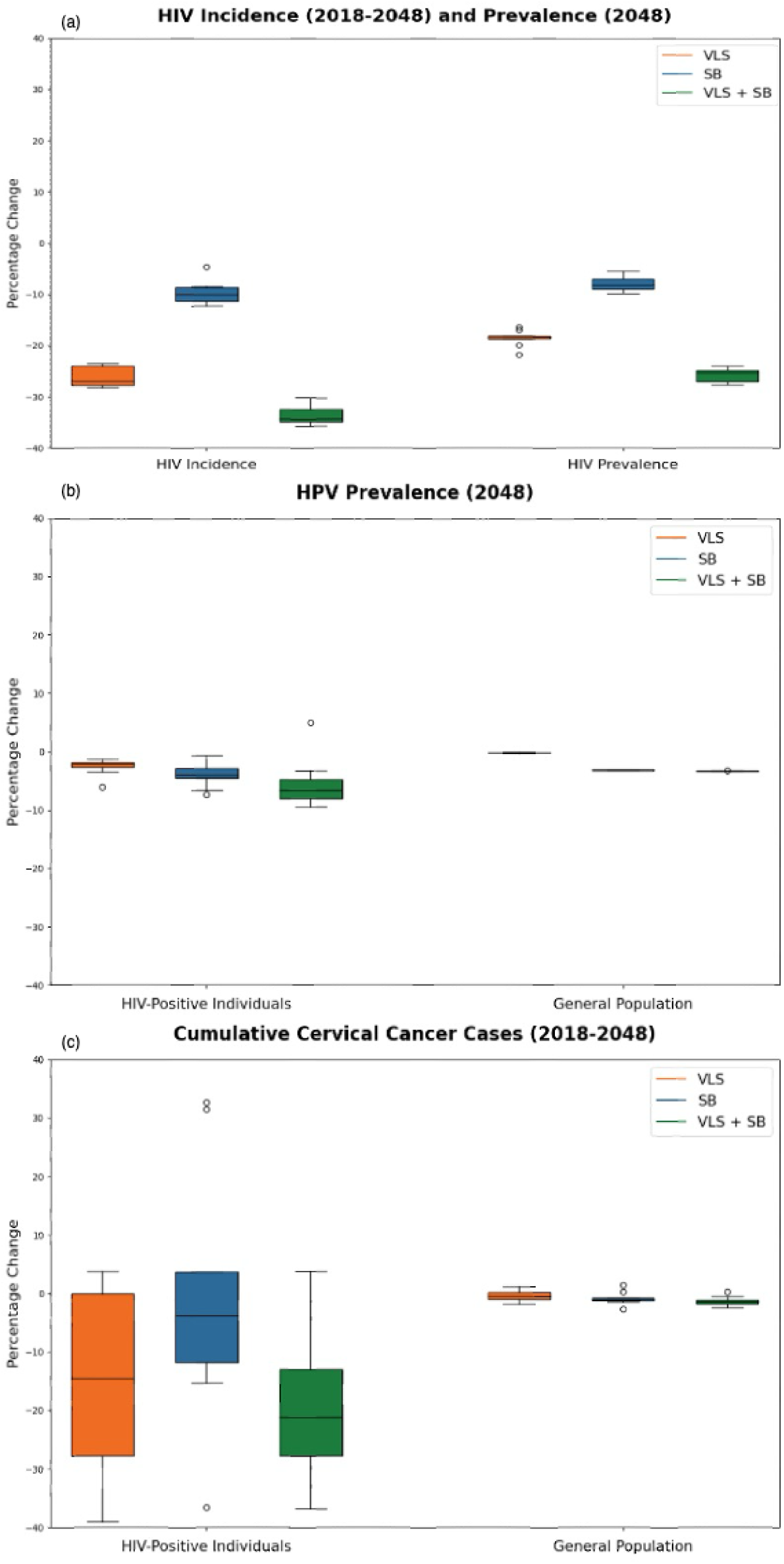
Percentage Change in Key Health Metrics Among Women Compared to ‘Baseline’ Under Three Interventions (VLS, SB, and VLS+SB): (a) Cumulative HIV incidence from 2018 to 2048 (left), HIV prevalence in 2048 (right).(b) HPV prevalence among the HIV-positive population in 2048 (left), HPV prevalence among the general population in 2048 (right).(c) Cumulative cervical cancer cases among the HIV-positive population from 2018 to 2048 (left), cumulative cervical cancer cases among the general population from 2018 to 2048 (right).*Note:* VLS = HIV care behavior intervention; SB = sexual behavior intervention; VLS+SB = combination of HIV care behavior intervention and sexual behavior intervention.

Among HIV-positive women, there was a 2.0% (range: 1.2% to 6.0%) decrease in HPV prevalence (Figure 1a) and a cumulative 14.5% (range: 3.9% to 39.9%) decrease in cervical cancer incidence (Figure 1c) because of indirect impact of VLS. This reduction results from improved VLS, which leads to stable CD4 counts, reducing the biological risk of HPV infection and increasing the likelihood of clearing HPV infections.

These results highlight the potential of addressing social needs to improve health outcomes among HIV-positive individuals and emphasize the importance of integrated intervention strategies.

### Sexual Behavior Intervention

Aligning the sexual behaviors—specifically the number of lifetime partners and condom use—of individuals engaged in exchange sex with those not engaged led to health improvements in our 30-year simulation. With this intervention, HIV prevalence decreased by 8.1% (range: 5.5% to 9.8%), and HIV incidence decreased by 10.0% (range: 4.6% to 12.2%) from 2018 to 2048 (Figure 1a).

Among HIV-positive women, HPV prevalence decreased by 4.4% (range: 2.8% to 6.5%) (Figure 1b). Specifically, HPV prevalence among those engaged in exchange sex decreased by 36.1% (range: 26.1% to 40.4%), while it changed by –1.4% (range: –4.6% to 2.1%) among those not engaged (Table 1). Among HIV-negative women engaged in exchange sex, HPV prevalence decreased by 32.8% (range: 32.7% to 33.0%), leading to an overall drop of 33.0% (range: 32.9% to 33.2%) in the exchange sex group (Table 1). Due to network effects, the entire population experienced a 3.3% (range: 3.2% to 3.4%) decrease in HPV prevalence.

**Table 1.**
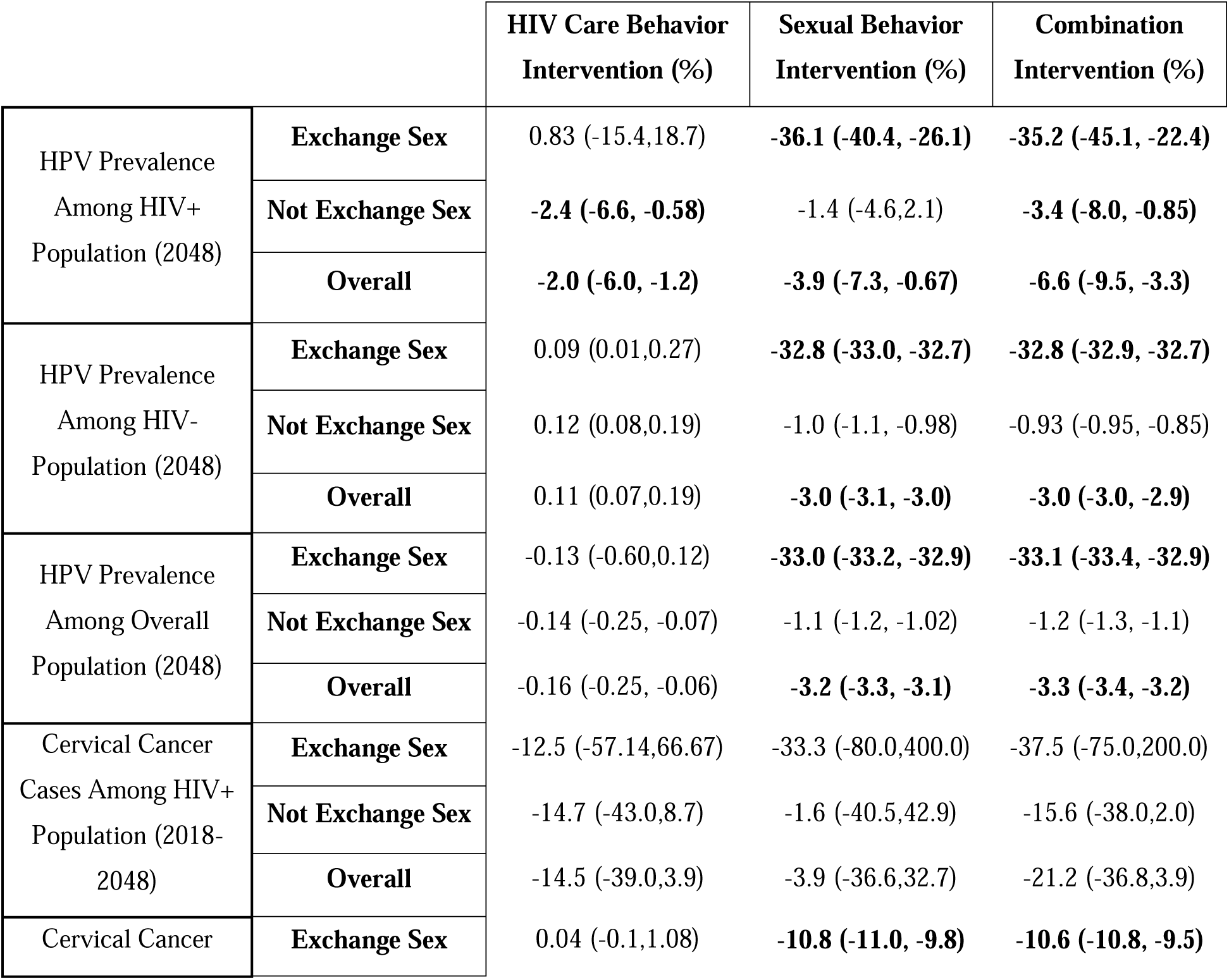
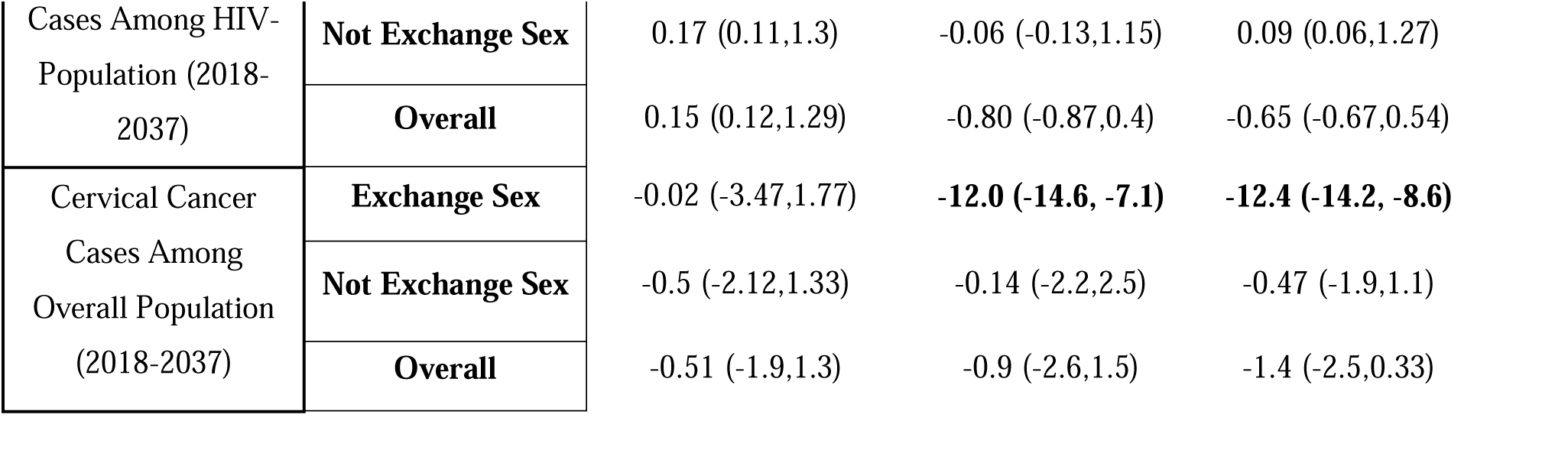
Percentage Change in Key Metrics Under Three Interventions Stratified by Exchange Sex Status.

Cervical cancer cases among HIV-negative individuals engaged in exchange sex decreased by 10.8% (range: 9.8% to 11.0%) (Table 1). Among HIV-positive individuals, cases showed a median decrease of 33.3% (range: –80.0% to 400.0%) (Table 1). The wide range of outcomes, spanning both positive and negative changes, can be attributed to the small proportion of the exchange sex group (approximately 10% among the HIV-positive population by the end of 2048) and the inherent randomness in individual case occurrences.

These results demonstrate that interventions addressing social needs among exchange sex such that their sexual behavior aligns with those of not exchange sex, the intervention demonstrates potential to reduce HIV and HPV prevalence across various populations and cervical cancer cases among exchange sex group.

### Combination of HIV Care Behavior and Sexual Behavior Intervention

By 2048, the combination intervention significantly reduced HIV prevalence and incidence across all population groups. HIV prevalence decreased by 25.3% (range: 24.0% to 27.6%) (Figure 1a), and HIV incidence from 2018 to 2048 reduced by 34.3% (range: 30.3% to 35.7%) (Figure 1a).

The intervention also substantially impacted HPV prevalence. Among HIV-positive individuals engaged in exchange sex, HPV prevalence decreased by 35.2% (range: 22.4% to 45.1%) (Table 1). In HIV-negative individuals engaged in exchange sex, HPV prevalence declined by 32.8% (range: 32.7% to 32.9%). The overall population experienced a 3.3% (range: 3.2% to 3.4%) reduction in HPV prevalence.

Among HIV-negative individuals engaged in exchange sex, cervical cancer cases decreased by 10.6% (range: 9.5% to 10.8%). Cervical cancer cases among HIV-positive individuals engaged in exchange sex showed a median change of –37.5% (range: –75% to 200%). For HIV-positive individuals not engaged in exchange sex, cases varied from –38.0% to 2.0%. The overall population saw a median change of –1.4% (range: –2.4% to 0.3%) (Table 1).

These results demonstrate that combined interventions effectively reduce HIV and HPV prevalence and decrease cervical cancer cases, particularly in high-risk groups. However, the variability in outcomes among HIV-positive individuals suggests the need for cautious adoption of these interventions and further research to address the observed disparities.

### Time Trend in Cumulative Cancer Cases

Figure 2 illustrates the annual percentage change in cumulative cancer cases among the HIV-positive population from 2007, showing median and range values. All three interventions—the HIV care behavior intervention (VLS), sexual behavior intervention (SB), and their combination (VLS+SB)—initially show an increase in cancer cases, peaking around the 2020s. This is followed by stabilization leading to a negative percentage change. The stabilization point is similar for both the HIV care behavior intervention and the combination intervention, while the sexual behavior intervention demonstrates a more modest change. Variability is highest around the peak period, indicating greater uncertainty then; over time, the range narrows, suggesting more consistent outcomes. Due to the small sample size, we calculated cumulative cancer cases up to 2048 to estimate long-term impact and mitigate randomness.

**Figure 2.**
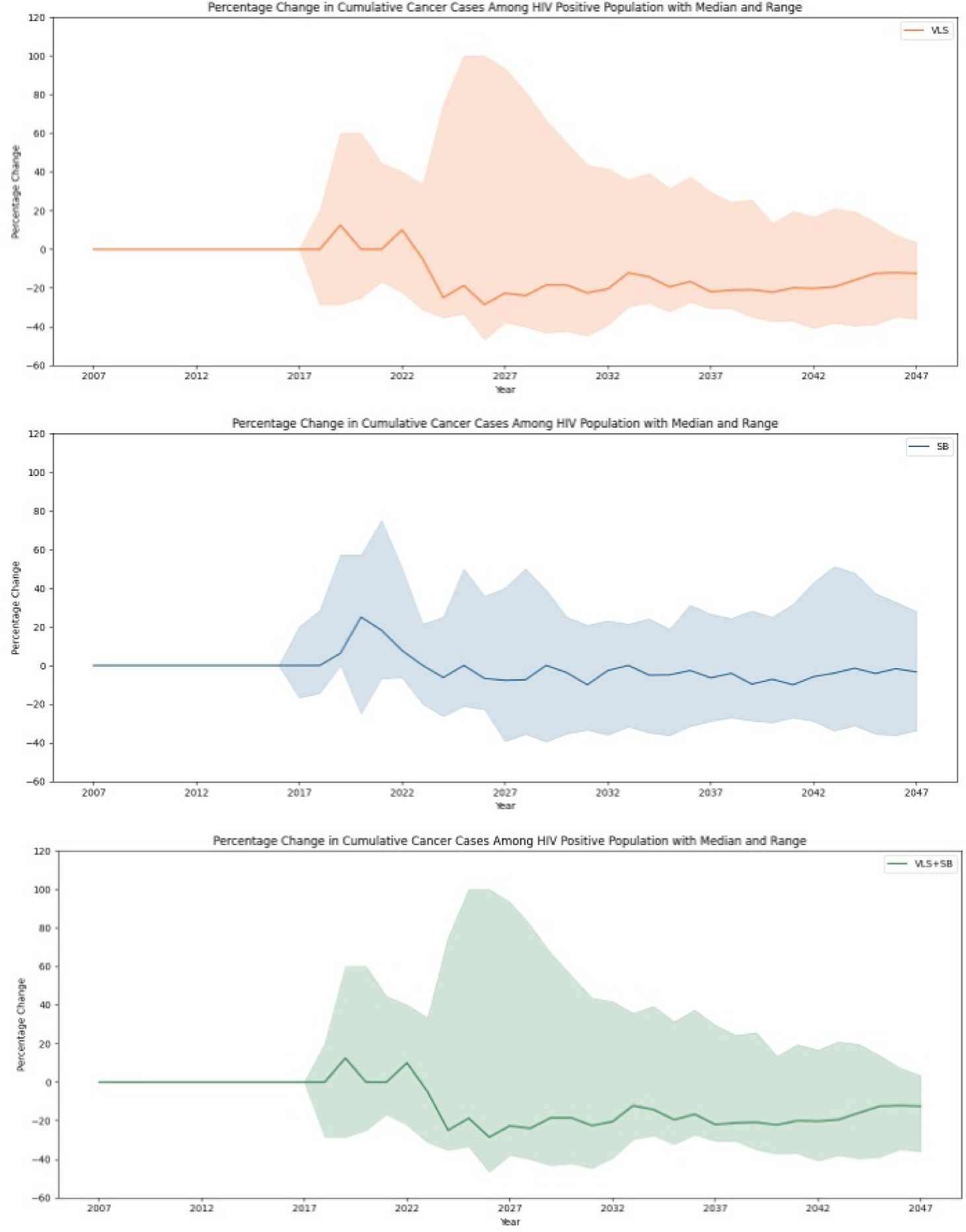
Percentage Change in Cumulative Cancer Cases among HIV Positive Population Under Three Interventions. Note: VLS = HIV care behavior intervention; SB = sexual behavior intervention; VLS+SB = combination of HIV care behavior intervention and sexual behavior intervention.

## 4. Discussions

In this study, we extended a recently developed MAC-HIV-CC model to include social conditions, enabling us to simulate the effects of behavioral and structural interventions on HIV, HPV, and cervical cancer among women. We explored hypothetical scenarios where addressing the social needs of socially disadvantaged groups could improve HIV care behaviors and sexual practices, and examined their impact on HIV, HPV and cervical cancer among women.

As part of validating our model’s projections for cervical cancer incidence, we reviewed similar modeling studies on the projection of cervical cancer cases in the U.S. under status quo interventions. Notably, two existing models ^61^ estimated that cervical cancer incidence would drop below 4 per 100,000 persons by 2038 and 2046, respectively. In comparison, our model projects this decline to occur around 2044–2045, demonstrating consistency with existing studies while offering an independent projection that aligns closely with available data.

Our findings show that addressing social needs in persons with HIV to improve VLS, significantly impacts disease outcomes: upto 18.3% reduction in HIV prevalence, 26.9% decrease in HIV incidence, a 2.0% reduction in HPV prevalence among HIV-positive women, and 14.5% decline in cervical cancer cases.

Sexual behavioral changes in persons who exchange sex reduced disease transmissions and improved health outcomes. The sexual behavior intervention resulted in an 8.1% reduction in HIV prevalence, a 10.0% decrease in HIV incidence, a 36.1% reduction in HPV prevalence among HIV-positive individuals engaged in exchange sex, and a 33.3% decrease in cervical cancer cases in the same group.

Combination interventions were even more effective, achieving a 25.3% reduction in HIV prevalence and a 34.3% decrease in HIV incidence. HPV prevalence and cervical cancer cases among HIV-positive individuals engaged in exchange sex decreased by 35.2% and 37.5%, respectively.

Unlike other modeling studies ^34,37^, which primarily estimate the impact of scaling up ART to increase the proportion of individuals with VLS, our study investigates the impact of targeting social needs as a way to increase VLS. By identifying where to focus these interventions, we provide a framework for addressing structural and behavioral factors that influence health outcomes, offering a more nuanced approach to public health strategy.

However, our results should be interpreted with caution due to several limitations. One limitation is that our model did not consider the full spectrum of social needs and restricted to individuals with HIV or those who engage in exchange sex. Our primary aim was to demonstrate a model framework that can evaluate interventions influencing multiple disease outcomes with shared transmission routes and risk factors. The accuracy of our model depends heavily on input data and assumptions. For example, sensitivity analyses could explore variations in diagnosis delay times and drop-out rates, affecting how quickly viral load suppression increases. ^62^.

In addition, we assumed interventions are 100% effective in closing the gaps in HIV care behaviors and sexual practices. Current clinical trials vary significantly in the social conditions, age and races they incorporate ^41,63,64^, making interpretation challenging. In the absence of expansive data, which can be challenging to gather, novel methods that infer effectiveness through ongoing program data could help address this gap.

We also treated HIV care interventions and sexual behavior interventions as independent in our combined analysis. However, relationships likely exist between exchange sex and other social conditions in our study.^65,66^. Future analyses should revisit this as more data become available.

Lastly, while the interventions generally showed decrease in most disease outcomes, variability in our results—especially in cancer cases among HIV-positive women—suggests caution. This variability may be due to small sample sizes. Future studies should use larger samples to enhance the reliability and robustness of these findings.

Despite these limitations, our study offers valuable insights into the significant role for use of mathematical models for structural intervention analyses. By integrating social conditions into the MAC-HIV-CC model, we have demonstrated the potential of behavioral and structural interventions to reduce HIV and HPV prevalence and decrease cervical cancer cases among women living with HIV. This underscores the importance of considering social determinants in public health strategies aimed at mitigating these diseases.

In conclusion, our study shows that jointly simulating social conditions and multiple interrelated diseases effectively evaluates the impact of interventions on HIV, HPV, and cervical cancer in the U.S. Addressing social needs among disadvantaged populations leads to reductions in disease prevalence and improves health outcomes through enhanced behaviors. These findings highlight the necessity of comprehensive intervention strategies that integrate structural and biological components to enhance public health outcomes and reduce health disparities. Our work provides a versatile framework for evaluating the cost-effectiveness of structural interventions across various health contexts and can guide future research and data collection efforts to develop more precise and impactful public health strategies.

## Data Availability

All data generated or analyzed during this study are included in this published article, its companion articles ^10,50^, and the supplementary information files.

