## Appendix for "A Modeling Framework for Evaluating the Synergistic Impact of Structural Interventions on Related Diseases: HIV and Cervical Cancer as Case Study"

**Supplemental Materials**

**Table S1 Associations between social conditions among HIV positive population and HIV care behavior.**

| **Pairwise Variables** | **Relative Risk Equation** | **Relative Risk Value** | **Source** |
| --- | --- | --- | --- |
| VLS, depression | Pr (not in VLS \| depression) / Pr (not in VLS\| not depression) | 1.56 | ^1^ |
| VLS, neighborhood | Pr(not VLS\| resident of census tract with > 20% in poverty) / Pr(not in VLS \| not a resident with > 20% in poverty) | 1.12 | ^2^ |
| VLS, housing | Pr(not in VLS \| homeless) / Pr(not in VLS \| housed) | 1.65 (1.39-2.38) | ^2–5^ |
| VLS, poverty | Pr(not in VLS\| at or below federal poverty level) / Pr(not in VLS\| above poverty level) | 1.49 | ^3^ |
| VLS, education | Pr(not in VLS\| no high school degree)/ Pr(not in VLS\| with high school degree or higher) | 1.3 | ^3^ |
| VLS, insurance | Pr(not in VLS\| uninsured)/ Pr(not in VLS\| insured) | 2.53 (1.12-3.41) | ^2–4^ |
| Education, housing | Pr(no high school degree\| \| homeless) / Pr(no high school degree\| housed) | 1.5 | ^6^ |
| Employment, housing | Pr(unemployed \| homeless)/ Pr(unemployed\| housed) | 1.41 | ^5,6^ |
| Housing, insurance | Pr(homeless \| not insured) / Pr(homeless \| insured) | 1.59 | ^3^ |
| Depression, housing | Pr(mental illness = positive screen for any diagnosis \| homeless)/ Pr(mental illness = positive screen for any diagnosis \| housed) | 1.28 | ^7^ |

**Table S2 Marginal distribution of social conditions among diagnosed HIV positive and HIV care behavior.**

| **Variable: X** | **X = 1 Description** | **Marginal Pr(X = 1)** | **Source** |
| --- | --- | --- | --- |
| Depression | Yes | 0.27 | ^8^ |
| Neighborhood | Resident of census tract with >= 18% in poverty | 0.38 | ^9^ |
| Housing | Homeless | 0.1 | ^10^ |
| Poverty | Below federal poverty level | 0.43 | ^10^ |
| Education | No high school degree | 0.17 | ^10^ |
| Insurance | None (public, Ryan White, or other) | 0.03 | ^3^ |
| Employment | Unemployed | 0.41 | ^10^ |

**Table S3 Marginal distribution of exchange sex and sexual behavior and their associations**

| **Variable: X** | **X = 1 Description / Relative Risk Equation** | **Marginal Pr(X = 1)** | **Source** |
| --- | --- | --- | --- |
| Exchange sex | Yes | 0.071 | ^11^ |
| Condom use | Varies by age and transmission group | | ^12^ |
| Number of partners | Power-law distribution, varies by transmission group | | ^12^ |
| **Pairwise Variables** | **Relative Risk Equation** | **Relative Risk Value** | **Source** |
| Condom use, exchange sex | Pr(no condom use \| exchange sex) / Pr(no condom use \| exchange sex) | 1.1 for HETF 1.2 for MSM | ^11,13^ |
| Number of partners, exchange sex | E[number of partners\| exchange sex] / E[number of partners\| not exchange sex] | 2.26 for both HETF and MSM | ^11^ |
